# Evaluating the Influence of Anti-Thyroid Antibodies on Thyroid Function in Newly Identified Hypothyroid Patients in Bangladesh

**DOI:** 10.64898/2025.12.15.25342318

**Authors:** Mohammad Abdul Hannan, Shahjada Selim, A SM Mesbah Uddin, Md. Masud Rana

## Abstract

**Background:** Millions of people worldwide suffer from thyroid dysfunction, and especially hypothyroidism, which is a prevalent endocrine disorder contributing extensively to systemic and metabolic illness. In hypothyroidism, triiodothyronine (T3) and thyroxine (T4), thyroid hormones that control metabolism in several organ systems, are insufficiently secreted.

**Objectives:** The objective of this study was to determine the effect of anti-thyroid antibodies on thyroid function in Bangladeshi newly diagnosed patients with hypothyroidism.

**Methods:** A cross-sectional analysis of adult patients with newly diagnosed hypothyroidism was carried out. Thyroid function tests (FT4, TSH), thyroid autoantibodies (anti-TPO, anti-Tg), symptoms, physical findings, and demographics were obtained and analyzed.

**Results:** The average age of the study participants was 36.07±11.00 years, and 70.1% were female. 72.7% of the cases were rural. 89% of the patients were antibody-positive, 81.8% anti-TPO, 55.2% anti-Tg, and 48.1% both. Enlargement of the thyroid (p<0.001) and gain of weight (p<0.043) were associated with antibody positivity. Grade 1 goitre alone was highly predictive of antibody positivity (AOR 11.766, p<0.001). Neither FT4 nor TSH correlated significantly with antibody titers. A significant correlation, however, was noted between anti-Tg and anti-TPO titers.

**Conclusion:** Recently developed hypothyroid patients usually have a condition named especially anti-TPO positive, and it is usually accompanied by goitre and family history. Even if the thyroid function tests are not conclusive, early diagnosis and better understanding of the disease process can be made by screening for thyroid antibodies.

## Introduction

Millions of people worldwide suffer from thyroid disorders, especially hypothyroidism, a prevalent endocrine condition that is primarily accountable for systemic and metabolic abnormalities. Triiodothyronine (T3) and thyroxine (T4) are thyroid hormones that govern metabolism in most organ systems and are released in deficient amounts in the event of hypothyroidism [1]. Prevalence differs at every site; iodine-deficient and autoimmune-susceptible populations have increased rates. Around 5% of the world’s population suffer from hypothyroidism, and a further 5% to 10% may suffer from subclinical hypothyroidism; females are overrepresented [2,3].

In regions where iodine is not deficient, autoimmune thyroid disease (AITD), encompassing atrophic thyroiditis and Hashimoto’s thyroiditis, is the most common cause of hypothyroidism [4]. The presence of circulating anti-thyroid antibodies, i.e., anti-thyroid peroxidase (anti-TPO) and anti-thyroglobulin (anti-Tg) antibodies, is a feature of AITD. In addition to their application as markers of diagnosis of AITD, these antibodies have been implicated in the pathophysiology of thyroid cell destruction by both humoral as well as cellular immunological mechanisms [5,6].

Over 90% of individuals with Hashimoto’s thyroiditis and up to 70% of subclinical hypothyroidism patients are positive for anti-TPO antibodies, which are anti-thyroid peroxidase, a critical enzyme used in the synthesis of thyroid hormones [7].

40–70% of AITD present with anti-Tg antibodies, which are anti-thyroglobulin, a precursor protein for T3 and T4, and commonly accompany anti-TPO antibodies [8]. The occurrence of these antibodies can be utilized to differentiate autoimmune hypothyroidism from other etiologies, like iodine deficiency, post-operative hypothyroidism, or drug-induced thyroid illness, and is a sure indication of autoimmune aetiology [9]. Anti-thyroid antibodies are not only of clinical significance more than just diagnosis. Their detection in patients with subclinical hypothyroidism has been reported by studies to have predictive value for overt hypothyroidism [10,11]. Long-term therapeutic decisions are also influenced by the fact that high anti-TPO titers are associated with greater gland damage and resistance to treatment with thyroxine [12]. Anti-thyroid antibodies are also linked with a higher risk of developing other autoimmune diseases such as rheumatoid arthritis, systemic lupus erythematosus, and type 1 diabetes [13].

Thyroid illness is common but frequently undiagnosed within South Asian nations like Bangladesh because they have poor access to health services, low community awareness, and poor diagnosing skills within primary and secondary healthcare facilities [14]. Even though serum TSH and free T4 are the most frequently applied markers in clinical practice, the measurement of antibodies is not a part of routine practice, so the autoimmune cause of hypothyroid patients goes undetected. The autoimmune status of these patients is still poorly understood, despite some hospital-based studies from the urban districts of Bangladesh reporting an increase in the diagnosis of hypothyroidism, predominantly among middle-aged women [15,16].

It is crucial to have baseline information regarding the prevalence of anti-thyroid antibodies in newly diagnosed cases of hypothyroidism in the context of changing global pandemics of non-communicable diseases like thyroid and autoimmune diseases(17). Early diagnosis, customized therapy, and slowing of the course of the disease are all likely to be helped by this information. In addition, identification of high-risk groups for family screening and follow-up of related autoimmune disorders may largely depend on screening for antibodies(18).

In this study, newly diagnosed Bangladeshi hypothyroid patients will have their thyroid hormone levels tested for the presence and impact of anti-thyroid antibodies (anti-TPO and anti-Tg)(19). This study is aimed at facilitating evidence-based inclusion of antibody tests in protocols for treatment of thyroid conditions in Bangladesh by demarcating the autoimmune nature of hypothyroidism in diagnosis. While thyroid conditions are commonly diagnosed in Bangladesh, anti-thyroid antibodies are not necessarily tested routinely, particularly in the secondary and primary levels of care. Because of this reality, autoimmune hypothyroidism might not be accurately diagnosed, which contributes to suboptimal care for new patients with the disease. A knowledge of the prevalence and impact of anti-thyroid antibodies in people with newly diagnosed hypothyroidism can predict the course of the disease, guide personalized therapy protocols, and give insight into the underlying aetiology. Thus, the aim of this study was to determine the impact of anti-thyroid antibodies (anti-TPO and anti-Tg) on thyroid hormone levels in newly diagnosed Bangladeshi patients with hypothyroidism.

## Methodology

### Study Design

In order to assess the impact of anti-thyroid antibodies on thyroid function among patients with newly developed hypothyroidism, a cross-sectional observational study design was used.

### Study Setting

The research was carried out at Bangabandhu Sheikh Mujib Medical University (BSMMU), Dhaka, Bangladesh, in the Department of Endocrinology. As a tertiary care unit serving a heterogeneous group of patients, BSMMU offers a strong platform for examining thyroid disorders among various clinical and demographic profiles.

### Study Population

The formula to calculate the sample size is as follows:

n = z 2 pq / d2

Where the increase in prevalence reported in some studies, p = Proportion or Percentage of Prevalence = 30–40% (Hari KKVS et al., 2012).

= 100 – 40 = 60% q = (1–p)

z = standard normal distribution (z distribution) value at a selected significance level or confidence level (1.96 for a fixed 95% confidence level).

d = Margin of error for prevalence (20 percent of 40)

Thus, from the above formula, [(1.96) 2 x 40 x 60] = n [(20 x 40)/100]2 = [(20 x 40)/100] / (3.8 x 2400) 9220 / 60 = 153.06 (about 154),

Thus, the sample size of this research is 154.

### Inclusion Criteria

- Adults 18 years and older who are newly diagnosed with primary hypothyroidism, as defined by decreased free T4 concentrations and increased thyroid-stimulating hormone (TSH) concentrations.
- Those who have never been treated for thyroid dysfunction.
- Patients who are willing to adhere to study protocol and sign informed consent.

### Exclusion Criteria

- Patients who have central or secondary hypothyroidism (e.g., secondary to pituitary or hypothalamic disease).
- Patients with systemic diseases impacting thyroid hormone function, such as non-thyroidal sickness syndrome.
- Pregnant or breastfeeding women.
- Patients undergoing continuous levothyroxine therapy, radioactive iodine ablation, or thyroidectomy.
- Patients with rheumatoid arthritis or systemic lupus erythematosus as autoimmune comorbidities.

### Sample Size

The determination of sample size was sufficient power to establish statistically significant associations between thyroid function and levels of TPOAb/TgAb. There was 154 patients enrolled, assuming 80% power, a 95% confidence interval, and an estimated effect size of 0.3.

## Data Collection

**Clinical Evaluation:** taking a full medical history to record symptoms, duration of illness, and whether there is a family history of autoimmune or thyroid disease. Physical examination, including palpation of the thyroid gland to feel for goitre or nodules.

### Laboratory Investigations

#### Thyroid Function Tests

Chemiluminescent immunoassay (CLIA) is utilized to determine the concentration of serum TSH, free T4, and free T3.

#### Autoantibody Quantities

TPOAb and TgAb were measured using enzyme-linked immunosorbent assay (ELISA).

#### Sociodemographic and Lifestyle Determinants

Systematic questionnaires was used to collect information on age, sex, BMI, dietary iodine consumption, smoking status, and access to healthcare.

#### Symptom Severity Scoring

To assess the intensity and impact of hypothyroidism symptoms, patients was filling out a validated symptom questionnaire.

#### Data Analysis: Descriptive Statistics

Means, medians, and frequencies was used to summarise the biochemical data, clinical aspects, and patient characteristics. TPOAb/TgAb values was used to stratify patients into low, moderate, and high subgroups. ANOVA or Kruskal-Wallis tests was used to evaluate group differences in clinical and biochemical data. The independent effects of TPOAb/TgAb levels on thyroid function was ascertained using multivariate linear regression models, which was account for variables such age, sex, and body mass index.

#### Ethical approval

This study was received Institutional Review Board (IRB) from BSMMU was provide ethical approval. All participants were asked to provide written informed consent. Patient data was anonymised in order to preserve data confidentiality.

## Results

Table 1 of the study presents the baseline characteristics of patients with newly diagnosed adult hypothyroidism. The average age is 36.07±11.00 years. The majority (70.1%) were female. Nine percent of the patients have a family history of thyroid disease. The prevalence of hypothyroidism was higher in rural areas (72.7%).

**Table 1:** Baseline characteristics of the newly detected adult hypothyroid patients.

| Variables | Frequency (n=154) |
| --- | --- |
| No. of study subjects | 154 (100.0%) |
| Age in years | $36.07 \pm 11.00$ |
| Gender |  |
| Male | 46 (29.9%) |
| Female | 108 (70.1%) |
| Family history of thyroid disease | 14 (9.1%) |
| Residence |  |
| Urban | 42 (27.3%) |
| Rural | 112 (72.7%) |

Table 2 here presents the baseline characteristics and antibody status among study participants. No difference in age was seen among the study population’s 89% positive for antibodies and 11% negative for antibodies. There were greater numbers of males and females who had positive antibodies, but results were not statistically significant. Positive antibodies were found in 86 percent of cases among hypothyroid patients with a positive family history of thyroid disease. Most of the hypothyroid subjects in rural and urban populations were positive for antibodies.

**Table 2:** Baseline characters and antibody status of the study subjects.

| Variables | Anti body |  | P value |
| --- | --- | --- | --- |
|  | Positive | Negative |  |
| No. of study subjects | 137 (89.0%) | 17 (11.0%) |  |
| Age in years (mean±SD) | 36.00 ± 11.01 | 36.64 ± 11.26 | 0.820* |
| Gender |  |  |  |
| Male | 39 (85.0%) | 7 (15.0%) | 0.280<sup>##</sup> |
| Female | 98 (91.0%) | 10 (9.0%) | 0.280<sup>##</sup> |
| Family history of thyroid disease | 12 (86.0%) | 2 (14.0%) | 0.684<sup>##</sup> |
| Residence |  |  |  |
| Urban | 38 (90.0%) | 4 (10.0%) | 0.483<sup>##</sup> |
| Rural | 99 (88.0%) | 13 (12.0%) |  |

Thyroid autoimmunity in hypothyroid patients is shown in figure 1. Anti-TPO antibodies were found in most newly diagnosed hypothyroid patients (81.8%). Anti-TG antibodies were found in 55.2% of hypothyroid individuals, and 48.1% had both antibodies.

**Figure 1:**
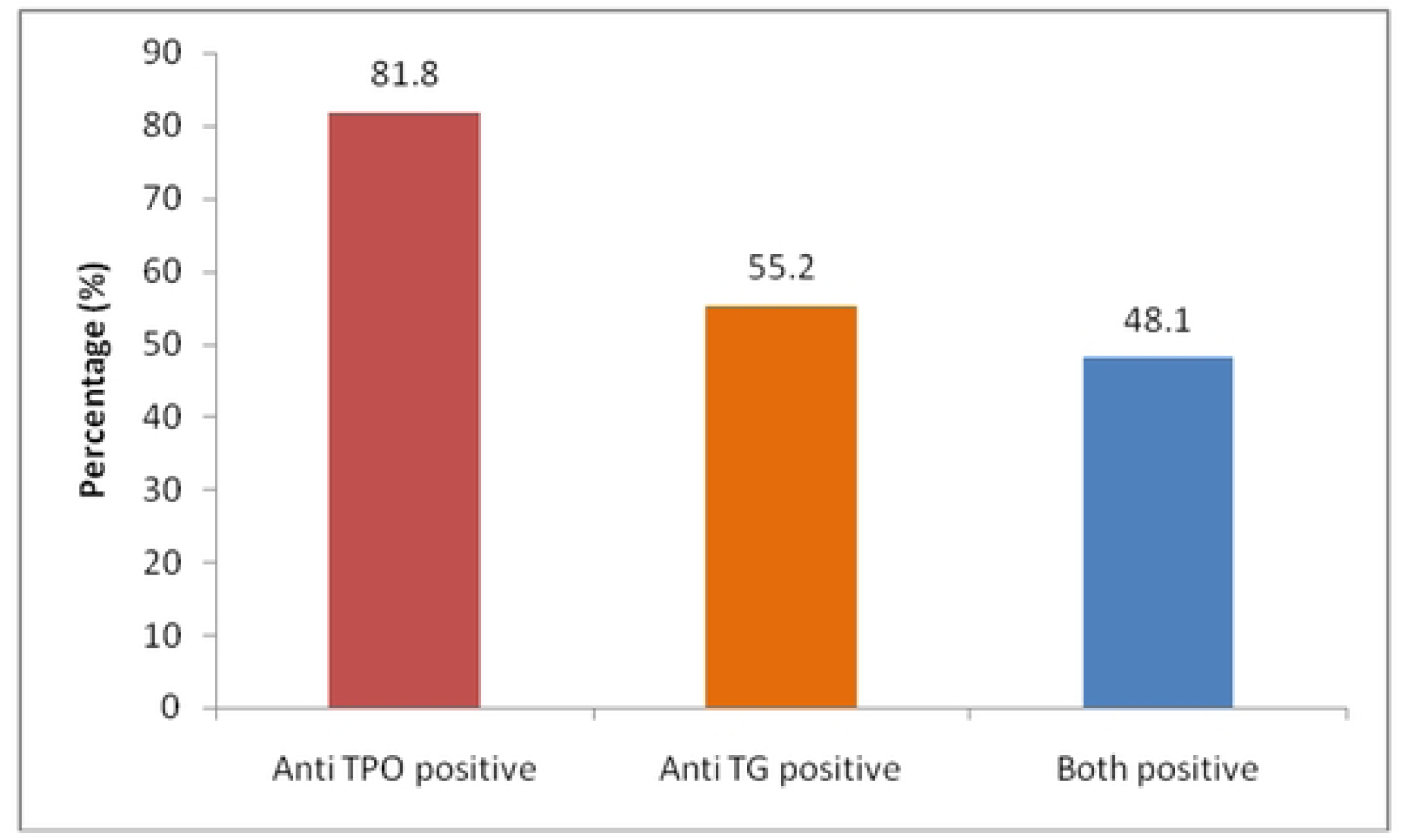
Frequency of presence of thyroid auto antibodies in study subjects.

Table 3 shows the presenting complaints of adult patients with hypothyroidism. 73.4% of the patients were overweight, and a much greater percentage of them had positive anti-thyroid antibodies (p = 0.043). Although more common among patients with positive antibodies, all other complaints were not statistically significant.

**Table 3:** Presenting symptoms of the newly detected adult hypothyroid patients.

| Presenting Symptoms | Total<br>n=154 | Anti body |  | P value |
| --- | --- | --- | --- | --- |
|  |  | Positive<br>(n=137) | Negative<br>(n=17) |  |
| Fatigability | 136 (88.3%) | 123 (90.4%) | 13 (9.6%) | 0.107<sup>#</sup> |
| **Weight gain** | 113 (73.4%) | 104 (92.0%) | 9 (8.0%) | **0.043<sup>##</sup>** |
| Decreased libido | 100 (64.9%) | 89 (89.0%) | 11 (11.0%) | 0.983<sup>##</sup> |
| Body ache | 96 (62.3%) | 85 (89.0%) | 11 (11.0%) | 0.831<sup>##</sup> |
| Voice change | 95 (61.7%) | 85 (89.0%) | 10 (11.0%) | 0.797<sup>##</sup> |
| Scalp hair loss | 85 (55.2%) | 75 (88.0%) | 10 (12.0%) | 0.750<sup>##</sup> |
| Menstrual disturbance | 84 (54.5%) | 76(90.0%) | 8(10.0%) | 0.511<sup>#</sup> |
| Cold intolerance | 82 (53.2%) | 73 (89.0%) | 9 (11.0%) | 0.979<sup>##</sup> |
| Constipation | 77 (50.0%) | 69 (90.0%) | 8 (10.0%) | 0.797<sup>##</sup> |
| Decreased sweating | 50 (32.5%) | 45 (90.0%) | 5 (10.0%) | 0.775<sup>##</sup> |
| Bad obstetrical history | 31 (20.1%) | 27(87.0%) | 4(13.0%) | 0.711<sup>#</sup> |
| Somnolence | 14 (9.1%) | 10 (71.0%) | 4 (29.0%) | 0.051<sup>#</sup> |
| Subfertility | 9 (5.8%) | 9 (100%) | 0 (0.00%) | 0.598<sup>#</sup> |
| Galactorrhoea | 1 (0.6%) | 1 (100.0%) | 0 (0.00%) | 1.000<sup>#</sup> |

Table 4 collates adult hypothyroid patients’ physical findings. BMI 26.44 ± 4.53, diastolic blood pressure (mmHg) 80 ± 8, systolic blood pressure (mmHg) 115 ± 14. 60.4% of the patients presented with thyroid enlargement, and a significantly greater number of them were anti-thyroid antibody-positive (p = 0.001). They were more antibody-positive but none of the other parameters was statistically significant.

**Table 4:**
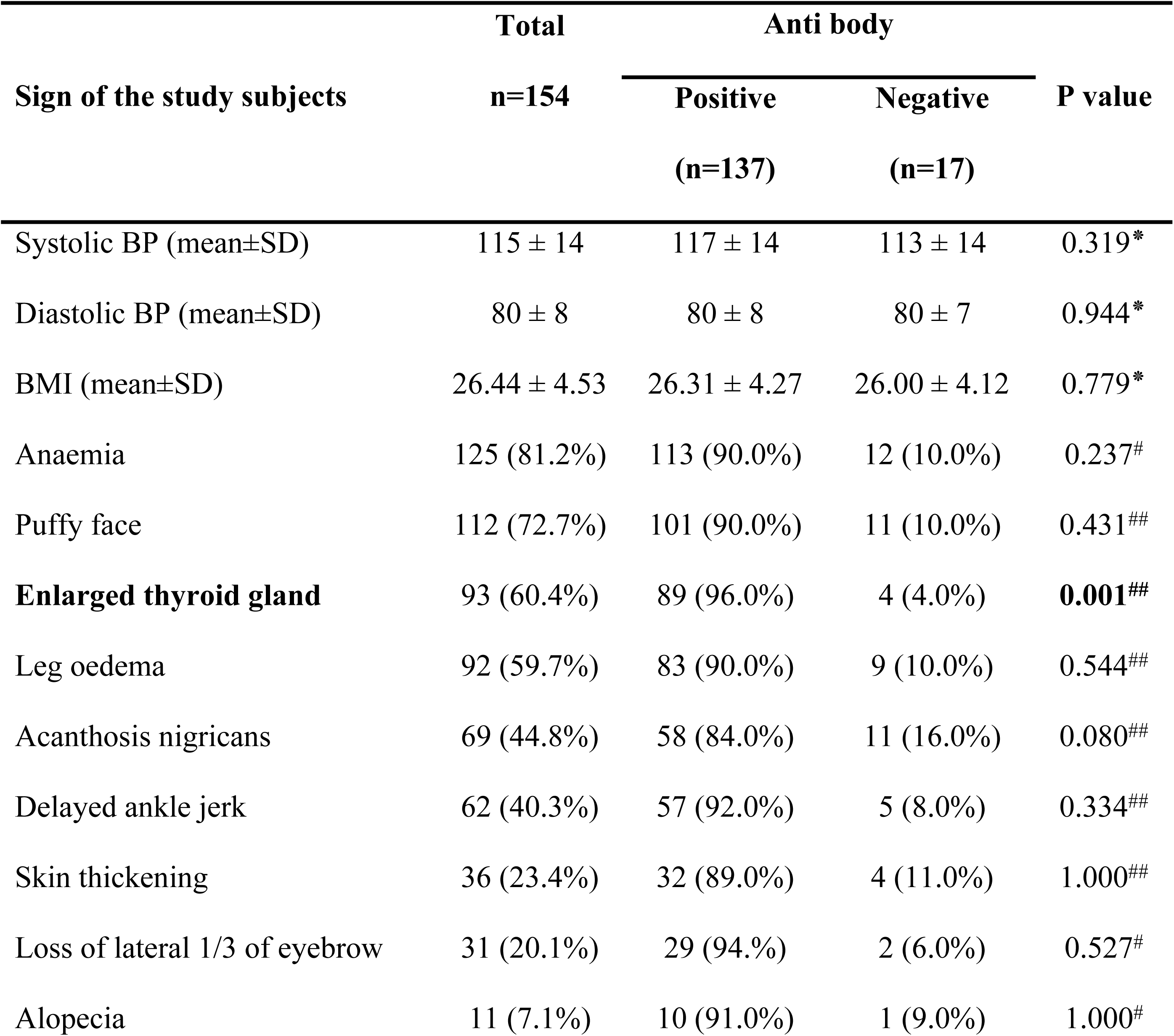
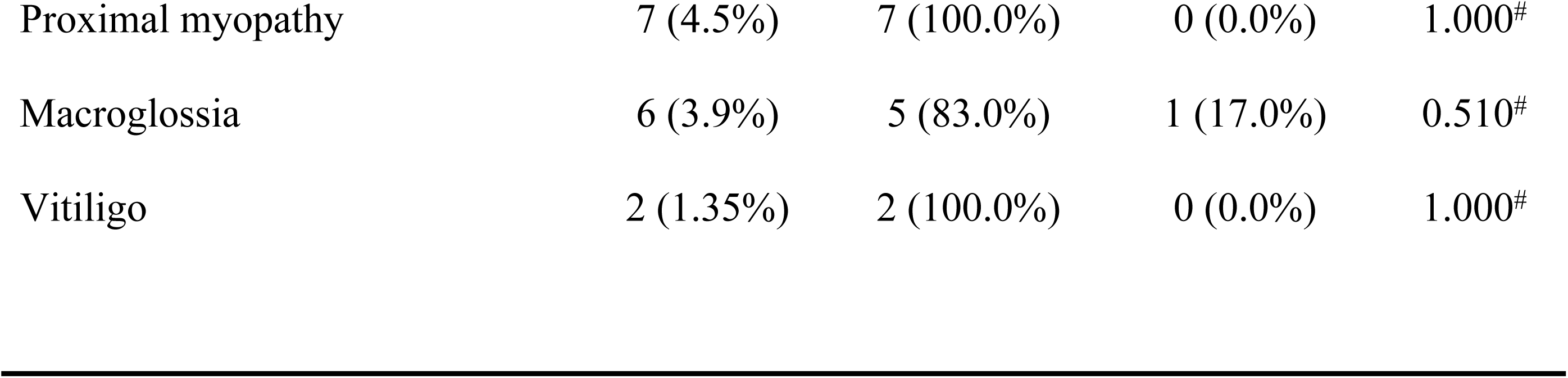
Physical findings of the adult newly detected hypothyroid patients (n=154)

| Sign of the study subjects | Total | Anti body |  | P value |
| --- | --- | --- | --- | --- |
|  | n=154 | Positive | Negative |  |
|  |  | (n=137) | (n=17) |  |
| Systolic BP (mean±SD) | 115 ± 14 | 117 ± 14 | 113 ± 14 | 0.319* |
| Diastolic BP (mean±SD) | 80 ± 8 | 80 ± 8 | 80 ± 7 | 0.944* |
| BMI (mean±SD) | 26.44 ± 4.53 | 26.31 ± 4.27 | 26.00 ± 4.12 | 0.779* |
| Anaemia | 125 (81.2%) | 113 (90.0%) | 12 (10.0%) | 0.237# |
| Puffy face | 112 (72.7%) | 101 (90.0%) | 11 (10.0%) | 0.431## |
| **Enlarged thyroid gland** | 93 (60.4%) | 89 (96.0%) | 4 (4.0%) | **0.001##** |
| Leg oedema | 92 (59.7%) | 83 (90.0%) | 9 (10.0%) | 0.544## |
| Acanthosis nigricans | 69 (44.8%) | 58 (84.0%) | 11 (16.0%) | 0.080## |
| Delayed ankle jerk | 62 (40.3%) | 57 (92.0%) | 5 (8.0%) | 0.334## |
| Skin thickening | 36 (23.4%) | 32 (89.0%) | 4 (11.0%) | 1.000## |
| Loss of lateral 1/3 of eyebrow | 31 (20.1%) | 29 (94.%) | 2 (6.0%) | 0.527# |
| Alopecia | 11 (7.1%) | 10 (91.0%) | 1 (9.0%) | 1.000# |
| Proximal myopathy | 7 (4.5%) | 7 (100.0%) | 0 (0.0%) | 1.000<sup>#</sup> |
| Macroglossia | 6 (3.9%) | 5 (83.0%) | 1 (17.0%) | 0.510<sup>#</sup> |
| Vitiligo | 2 (1.35%) | 2 (100.0%) | 0 (0.0%) | 1.000<sup>#</sup> |

Table 5 indicates antibody positivity with the grade of goitre and hypothyroidism. Increasing grade of goitre was observed in an increasing number of patients who were found to be antibody positive

**Table 5:** Presence of goiter and antibody positivity among study subjects.

| Goiter grade | Total<br>n=154 | Anti body |  | P value |
| --- | --- | --- | --- | --- |
|  |  | Positive | Negative |  |
|  |  | n=137 | n=17 |  |
| 0 grade | 23 (14.9%) | 15 (10.9%) | 8 (47.1%) | <0.001 |
| 1 grade | 41 (26.6%) | 36 (26.3%) | 5 (29.4%) | 0.999 |
| 2 grade | 90 (58.4%) | 86 (62.8%) | 4 (23.5%) | **0.003** |
| Total | 154 (100.0%) | 137 (100.0%) | 17 (100.0%) | <0.001 |

In the table 6, shows thyroid function status of the newly detected hypothyroid patients. No statistically significant influence of anti thyroid antibody upon FT4 and TSH was found.

**Table 6:**
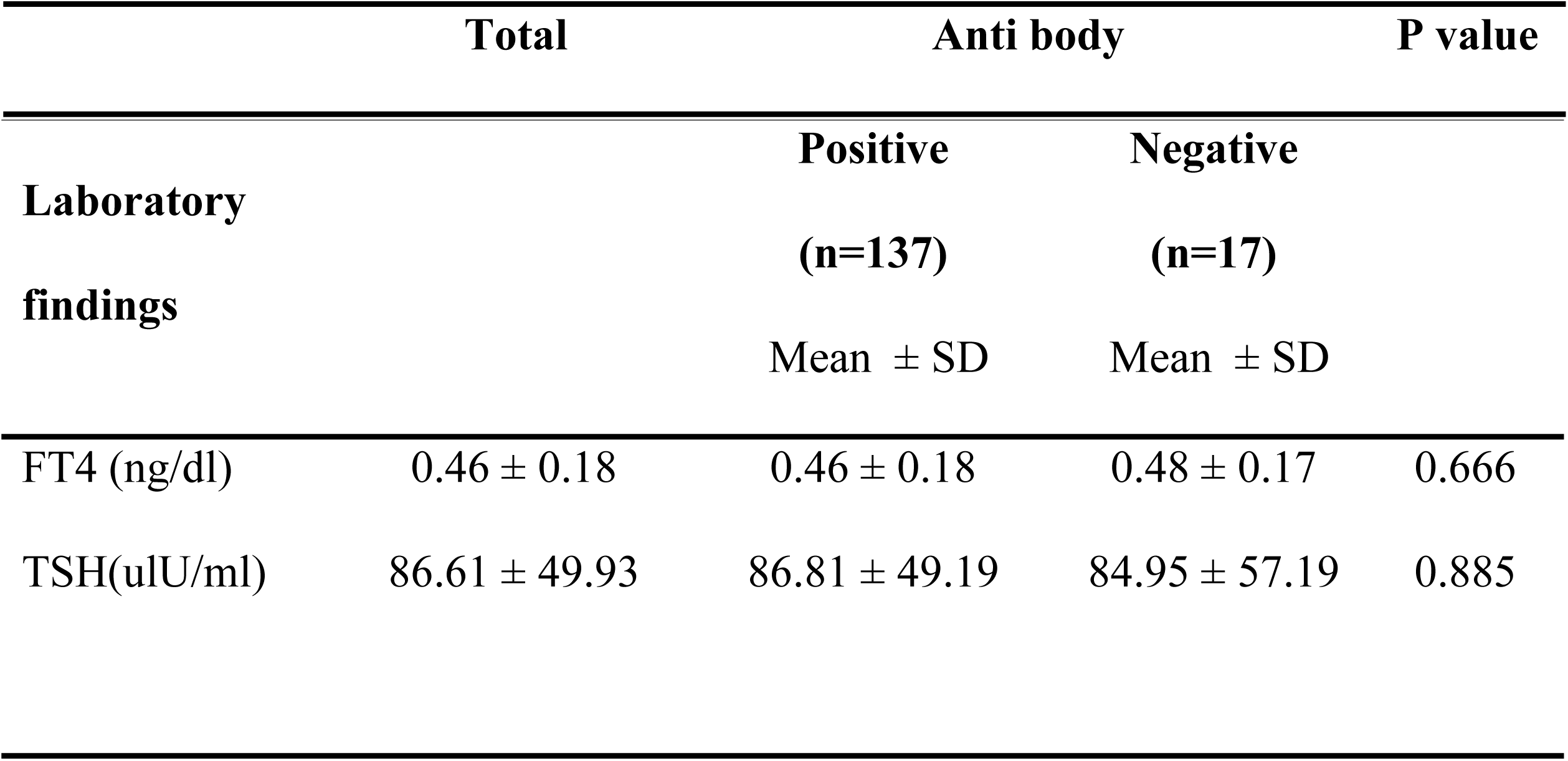
Thyroid function and autoimmune status of the newly detected hypothyroid patients.

In table 7, shows the Spearman correlation of serum anti thyroid antibodies with serum FT4 and TSH. None of the antibodies were significantly correlated with FT4 and TSH. But anti TPO titer vs anti Tg found statistical significant correlation

**Table 7:** Correlation of Anti-TPO and Anti-Tg with FT4 and TSH.

| Variables | r value | p value |
| --- | --- | --- |
| Anti-TPO titer vs. FT4 | -0.068 | 0.402 |
| Anti-TPO titer vs. TSH | 0.077 | 0.344 |
| Anti-Tg titer vs. FT4 | -0.158 | 0.050 |
| Anti-Tg titer vs. TSH | -0.037 | 0.650 |
| Combined Anti-TPO and Anti-Tg vs FT4 | 0.041 | 0.615 |
| Combined Anti-TPO and Anti-Tg vs TSH | -0.014 | 0.862 |
| Anti-TPO titer vs. anti-Tg | 0.198 | 0.014 |

Multiple logistic regression with anti-Tg and anti-TPO antibodies as independent variables is shown in Table 8. Antithyroid antibody positivity was found to be associated with grade 1 goitre (p <0.001, OR 11.766; 95% CI 2.992 – 46.266). It is 11.7 times more likely to be positive for thyroid antibody if there is grade 1 goitre. Anti-thyroid antibody positivity was not found to be associated with any other variable.

**Table 8:** Multiple regression analysis with anti-TPO and anti-Tg antibody as dependent variable.

| Variables | B | S.E. | Wald | p-value | OR | 95% CI of OR |  |
| --- | --- | --- | --- | --- | --- | --- | --- |
|  |  |  |  |  |  | Lower | Upper |
| Age | -0.005 | 0.025 | 0.049 | 0.825 | 0.995 | 0.947 | 1.044 |
| Sex (male) | 0.239 | 0.594 | 0.161 | 0.688 | 1.270 | 0.396 | 4.071 |
| Residence (rural) | -0.193 | 0.671 | 0.082 | 0.774 | 1.212 | 0.325 | 4.516 |
| Goiter (Grade 1) | 2.465 | 0.699 | 12.454 | 0.000 | 11.766 | 2.992 | 46.266 |
| Goiter Grade 2) | 1.148 | 0.721 | 2.538 | 0.111 | 3.153 | 0.768 | 12.952 |
| FT4 | 1.265 | 2.073 | 0.373 | 0.542 | 3.544 | 0.061 | 206.09 |
| TSH | -0.001 | 0.008 | 0.008 | 0.931 | 0.999 | 0.984 | 1.015 |

## Discussion

This study highlights the importance of thyroid autoimmunity by reporting extensive baseline features, clinical presentation, and serological profiles of adult patients newly diagnosed with hypothyroidism. The mean age of the patients was 36.07±11.00 years, and 70.1% were females. This concurs with global epidemiological patterns that report that women in childbearing ages are more susceptible to autoimmune thyroid disease and hypothyroidism (17,18). The rural area can account for the high prevalence of hypothyroidism due to disparity in access to healthcare, late diagnosis, or exposures to environmental substances affecting the thyroid function in the regions (19).

Autoantibodies against the thyroid were present in a high percentage (89%) of patients with hypothyroidism, which is suggestive of the autoimmune aetiology in most of these patients. The commonest antibody detected was anti-thyroid peroxidase (anti-TPO) followed by anti-thyroglobulin (anti-Tg) in 48.1% of patients. The results concur with other reports which found anti-TPO to be a more sensitive marker for autoimmune thyroiditis than anti-Tg (20,21). The genetic character of autoimmune thyroid disorders is also suggested from the findings that 9.1% were associated with a familial history, and 86% of them were antibody positive (22).

A more intense autoimmune process or a longer history of disease before diagnosis may be the explanation for the significantly higher prevalence of such symptoms as weight gain among antibody-positive persons, even in the absence of any significant differences in age or sex between antibody-positive and antibody-negative persons (23). Although not statistically significant, other symptoms also occurred more often among antibody-positive persons.

Goitre was detected on physical exam and was highly related to antibody positive. In addition, goitre grades larger were present in relation with higher antibody positivity. Grade 1 goitre was a strong clinical predictor of underlying thyroid autoimmunity, as evident from regression analysis, and also that goitre highly increased the odds of being antibody positive. These are consistent with earlier evidence that goitre usually occurs as autoimmune thyroiditis, especially in its early or subclinical presentation (24).

Thyroid function tests (TSH, FT4) were not significantly different between antibody-positive and -negative groups despite the fact that antibodies were strongly correlated with clinical and physical findings. Also, no such clear correlation between titers of antibodies and either FT4 or TSH was established using Spearman correlation. But one such interesting correlation between anti-TPO and anti-Tg titers demonstrates the fact that these antibodies can be raised simultaneously in certain autoimmune diseases (25, 26). Even if the outcome of thyroid function tests does not reflect the severity of the condition directly, in the current study, the diagnostic significance of anti-TPO and anti-Tg antibodies in hypothyroidism has been suggested, particularly in goitrous patients and those with familial history. In order to examine the aetiology of the condition and the risk of its development, in the current findings, the routine use of antibody screening in hypothyroid patients is recommended (26).

## Conclusion

This study found with the most common marker being anti-TPO antibody, the present study emphasizes the important role of thyroid autoimmunity in new onset adult hypothyroid patients. Clinical characteristics like weight gain, thyroid enlargement, and goitre grade were very much associated with antibody positive. Being grade 1 goitre was very highly raising the likelihood of antibody positivity irrespective of thyroid function tests (FT4, TSH) not differing significantly with antibody status. In order to make possible the early detection and specific treatment of autoimmune hypothyroidism, these results emphasize the value of periodical anti-thyroid antibody screening, particularly in those patients who have goitre or a family history of thyroid disease.

## Data Availability

After acceptimng the manuscript

## Acknowledgments

All participants who volunteered are appreciated by the study with thanks. Prior to the commencement of the study, written informed consent was obtained from each participant.

## Author contributions

SS and MAH had the conception and coordination of the study. MAH was responsible for data collection, critical assessment, validation, and visualization. ASMU contributed with a critical assessment and helpful comments to enhance the manuscript. MMR had the writing of the manuscript and data analysis. Each author reviewed, read, and approved the final version of the manuscript.

